# CIGB-258 immunomodulatory peptide: a novel promising treatment for critical and severe COVID-19 patients

**DOI:** 10.1101/2020.05.27.20110601

**Authors:** R Venegas-Rodriguez, R Santana-Sanchez, R Peña-Ruiz, M Bequet-Romero, M Hernandez-Cedeño, B Santiesteban-Licea, A Garcia, PR Aroche, D Oliva-Perez, LM Ortega-Gonzalez, J E Baldomero, LR Cruz, G Guillén, G Martinez-Donato, MC Dominguez-Horta, for the CIGB-258 Study Group

## Abstract

This study characterizes the first clinical application of CIGB-258 in COVID-19 patients. CIGB-258 is an immunoregulatory peptide, derived from the cellular heat shock protein 60 (HSP60). Sixteen patients with COVID-19 in serious (31%) or critical (69%) conditions were included in this study. All critically ill patients recovered from the respiratory distress condition. Two of these patients had a fatal outcome due to nosocomial infections.The five seriously ill patients considerably improved. C-reactive protein (CRP) and interleukin-6 (IL-6) levels significantly decreased during treatment. CIGB-258 seems to be an effective and safe treatment option in COVID-19 patients under cytokine storm.

TRIAL REGISTRATION: RPCEC00000313

## Introduction

Several authors have described that a subgroup of COVID-19 patients progresses towards a serious or critical stage, characterized by hyperinflammation (1). This phase is mediated by high levels of proinflammatory cytokines (2). During cytokine storms most patients arrive to cardiovascular collapse, multiple organ failure, and death (3).

In this context, approved therapies for autoimmune diseases are being used to control hyperinflammation and reduce mortality in patients with COVID-19. Treatments such as specific monoclonal antibodies for IL-1 (Anakinra), IL-6 (Tocilizumab) and Janus kinase inhibitors are being used (4). These treatments could reduce hyperinflamation but will undoubtedly cause immunosuppression. This is an unfavorable outcome for COVID-19 patients since virus-infected cells must eventually be cleared out by the immune system. Other approaches are being intensively assessed. Among them, CIGB-258 (previously called APL1 or CIGB-814 and hereafter CIGB-258) is an immunoregulatory peptide derived from the human heat shock protein 60 (HSP60). This protein increases its concentration during viral infections and inflammation. Peptides derived from HSP60 may represent danger signals that can trigger physiological inflammatory responses. However, peptides also derived from HSP60 can induce T cells with regulatory function. Therefore, different groups of researchers have assigned an immune response regulating role to HSP60 (5). CIGB-258 increases the frequency of regulatory T cells (Treg) and their suppressive capacity against the antigen responding effector CD4+T cells. Furthermore, this peptide induced a reduction of proinflammatory cytokines: TNFα, IL-17 and IFNg during preclinical studies (6,7) and phase I clinical trials (8).

Pharmacogenomics and proteomics of this peptide action indicate its ability to reduce inflammation mediators inducing neutrophil, macrophage and monocyte activation, hence promoting hyperinflammation (9).

Currently, a phase II clinical trial with this HSP60-derived immunoregulatory peptide is ongoing in patients with Rheumatoid Arthritis (RA), and so far 185 patients have been treated with this peptide with an important safety profile.

This evidence supports the use of CIGB-258 to control SARS-CoV-2 induced hyperinflammation. In this study, we report the therapeutical response of CIGB-258 in sixteen patients with COVID-19 in serious or critical conditions. Results indicate the preliminary evidence of the clinical efficacy of this peptide for the treatment of severe and critical COVID-19 patients.

## Methods

### Patients

Patients in the study were included from March 31 to April 22 2020 at the Luis Diaz Soto and Pedro Kouri Hospitals in Havana, Cuba. The Ethics and Scientific Committees of each study site and the Cuban Regulatory Authority (CECMED, http://www.cecmed.cu/) approved the protocol. Patients or their legal representatives signed the informed consent before using CIGB-258. All patient data were anonymously recorded to ensure confidentiality.

Patients with COVID-19 in seriously or critically ill conditions included in this study, were diagnosed according to the protocol of the Ministry of Public Health of the Republic of Cuba (http://infomed.sld.cu/anuncio/2020/05/11/ministerio-de-salud-publica-protocolo-de-actuacion-nacional-para-la-COVID-19). All patients received standard therapy according to the above cited protocol.This clinical trial was registered under number RPCEC00000313 at the Cuban Clinical Trial Registry (www.registroclinico.sld.cu).

### Procedures

Demographic characteristics and comorbidities of patients, treatments, laboratory results, chest x-ray and clinical outcome of the patients were obtained from the medical records. Serum samples were obtained before the treatment (T0) and at 24, 48, 72 and 96 hours. C reactive protein (CRP) was considered elevated when it was higher than 5.0 mg/L. Interleukin (IL)-6 was detected in neat serum samples using the Human CD8+ T-Cell Magnetic Bead Panel (HCD8MAG15K17PMX, EMD Millipore, Germany) according to the manufacturer’s instructions. Assay detection limits for IL-6 were between 0.91 and 1940 pg/mL. Results were acquired using the Luminex® analyzer (MAGPIX® EMD Millipore, Germany), and processed in the Milliplex Analyst software v 5.1.0.0 (EMD Millipore, Germany). The serum samples from fifteen healthy adults were used as controls for cross-comparisons.

### Safety

Data from patients were collected during the treatment according to Regulation 45/2007 from the Cuban Regulatory Authority: “Requirements for reporting adverse events in ongoing clinical trials, based on WHO regulations”. This regulation agrees with the “National Cancer Institute Common Toxicity Criteria Adverse Event version 3.0” (National Cancer Institute, Frederick, MD, USA).

### Statistical analysis

All patients receiving CIGB-258 were included in the clinical, radiological, laboratory and safety assessments. Adverse events (AEs), vital signs, laboratory tests, Chest X-ray and evidence of therapeutic effects were descriptively compared between baseline (T0) and data collected from patients after starting the CIGB-258 treatment with no formal statistical tests.

CRP and IL-6 data were analyzed using GraphPad Prism version 7.05 (GraphPadSofware, San Diego California, USA). Samples were examined for normality and equal variance with Kolmogorov-Smirnov and Bartlett’s tests, respectively. CRP levels were expressed as means, and their differences were analyzed with ANOVA and Tukey’s post hoc test. Kruskal Wallis and Dunn’s post hoc test were used for IL-6 analysis. P values less than 0.05 were considered statistically significant.

## Results

### Baseline characteristics and clinical description of patients

Sixteen patients with COVID-19 in serious or critical conditions were treated with CIGB-258. Demographic characteristics of patients, clinical classification and comorbidities are summarized in Table 1. Five (31%) patients were seriously ill, and eleven (69%) patients were critically ill. All critically ill patients were under invasive ventilation when starting the CIGB-258 treatment.

**Table 1:** Demographic characteristics of COVID-19 patients treated with CIGB-258 and results of the therapy

| Case No. | Sex | Clinical classification | Comorbidities | Respiratory distress at the end of treatment | Clinical outcomes |
| --- | --- | --- | --- | --- | --- |
| 1 | M | Critically ill | Cardiac arrhythmia | Recovered | Alive, discharged from Hospital |
| 2 | M | Critically ill | Hypertension<br>Diabetes<br>Obesity | Recovered | Alive, discharged from Hospital |
| 3 | M | Critically ill | None | Recovered | Alive, discharged from Hospital |
| 4 | F | Critically ill | Hypertension | Recovered | Alive, discharge from Hospital |
| 5^ | F | Critically ill | Adrenal gland tumor<br>Diabetes<br>Obesity<br>Cushing's syndrome,<br>Multiple thyroid nodules | Recovered | Alive, discharge from ICU |
| 6^ | M | Critically ill | Bladder cancer,<br>Diabetes | Recovered | Alive, discharged from Hospital |
| 7 | M | Critically ill | None | Recovered | Alive, discharged from Hospital |
| 8 | M | Critical ill | Hypertension<br>bronchial asthma<br>Ischemic heart disease | Recovered | Alive, discharged from Hospital |
| 9 | M | Critically ill | Hypertension,<br>Obesity,<br>Bronchial asthma | Recovered | Alive, discharged from Hospital |
| 10 | M | Seriously ill | None | Recovered | Alive, discharged from Hospital |
| 11 | M | Seriously ill | Hypertension<br>Diabetes<br>Ischemic heart disease | Recovered | Alive, discharged from Hospital |
| 12 | F | Seriously ill | Diabetes<br>Ischemic heart disease | Recovered | Alive, discharged from Hospital |
| 13* | M | Critically ill | bronchial asthma | Recovered | Deceased |
| 14* | F | Critically ill | Hypertension | Recovered | Deceased |
| 15* | F | Seriously ill | Glomerulopathy | Recovered | Alive, discharged from Hospital |
| 16 | M | Seriously ill | Hypertension | Recovered | Alive, discharged from Hospital |

\*: Treatment with monoclonal antibody Itolizumab (11).
^: Dose of CIGB-258: 2mg/12 hours M: male; F: Female; ICU: intensive care unit

Seriously ill patients had dyspnea, fever and fatigue. These patients were treated with oxygen therapy, including nasal cannula or oxygen mask.

The median age (m in-max) of patients was 61 (19-91) years old. Thirteen patients had one or more comorbidities, including: hypertension, diabetes, obesity, bronchial asthma, ischemic heart disease, and cancer.

### Therapy Outcomes

Nine critically ill patients were treated with 1mg of CIGB-258 every 12 hours and two patients were treated with 2 mg every 12 hours (Table 1).

All critically ill patients recovered from their respiratory distress condition and were extubated. They continued therapy with CIGB-258 for another 72 hours, after extubation. Unfortunately, two patients died from nosocomial infection. Eight of them were discharged from the hospital after an average of ten days under CIGB-258 treatment.

Seriously ill patients were treated with 1mg of CIGB-258 each 24 hours. These patients had a marked improvement in their clinical condition after 48 hours of treatment with CIGB-258. Fever disappeared and patients did not need further oxygen supplementation. These patients were discharged from the hospital after an average of seven days of treatment with CIGB-258.

The clinical and gasometric improvement was corroborated by radiology. Representative images of a patient before treatment, 48 hours after starting therapy, and after the extubation are shown in figure 1.

**Figure 1.**
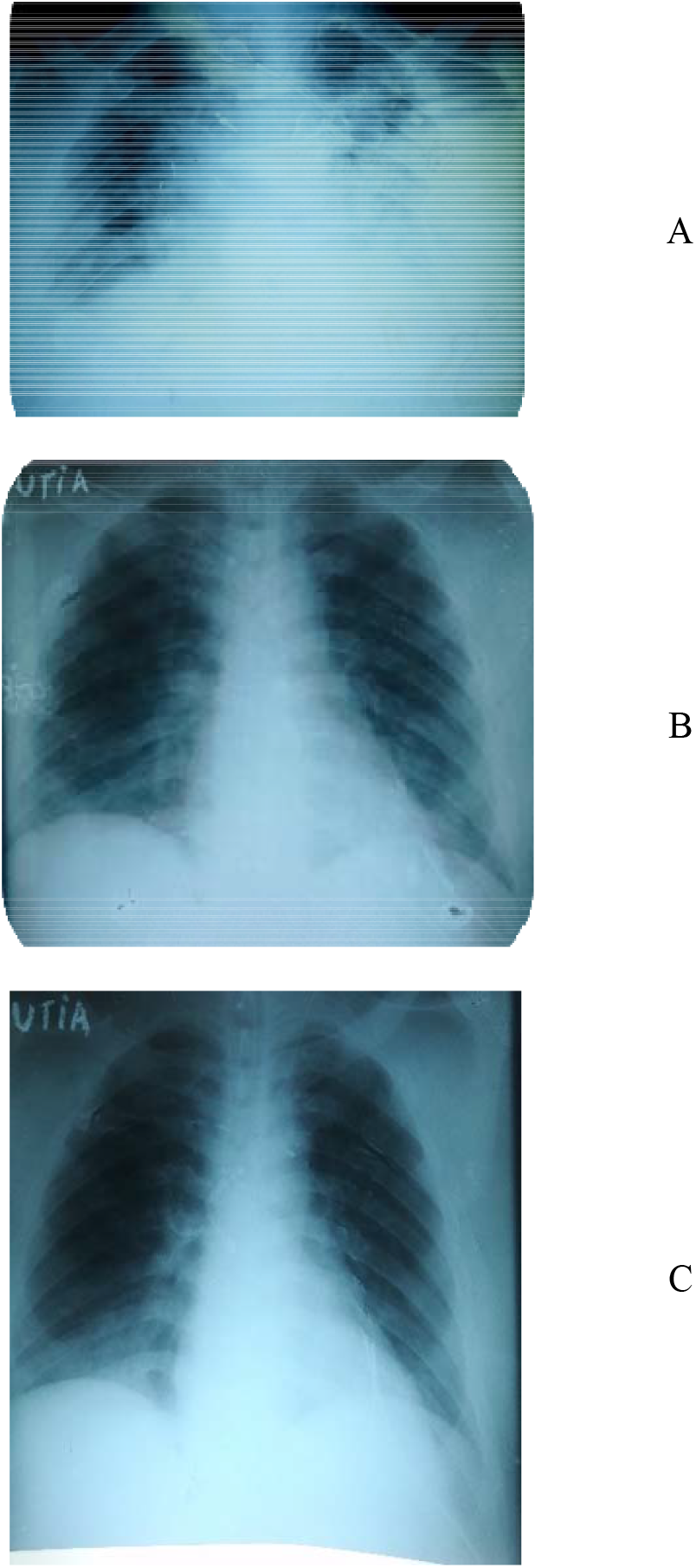
Progress of thorax X-ray imaging of acute respiratory distress syndrome in a representative patient. A: Thorax X-ray before mechanical ventilation: Opacity of both hemithorax. Right hemithorax with a heterogeneous appearance. Left hemithorax in mass with bilateral perihilar infiltrate. Widening of mediastinum with a vascular appearance. B: Thorax X-ray after 48 hours of CIGB-258 therapy: Increased bronchovascular markings. Small reticular lesions in the right lung base. Cardiothoracic ratio (CTR) in the upper normal limit C: Thorax X-ray before extubation: Improvement of thorax X-ray imaging with increased bronchovascular bundles

### Effect of CIGB-258 on Laboratory parameters

Laboratory parameters associated with hyperinflammation including liver enzymes, ferritin, D-dimer, fibrinogen and lactate dehydrogenase gradually normalized.

Specifically, CRP kinetics is shown in Figure 2. CRP levels decreased significantly during treatment with CIGB-258.

**Figure 2.**
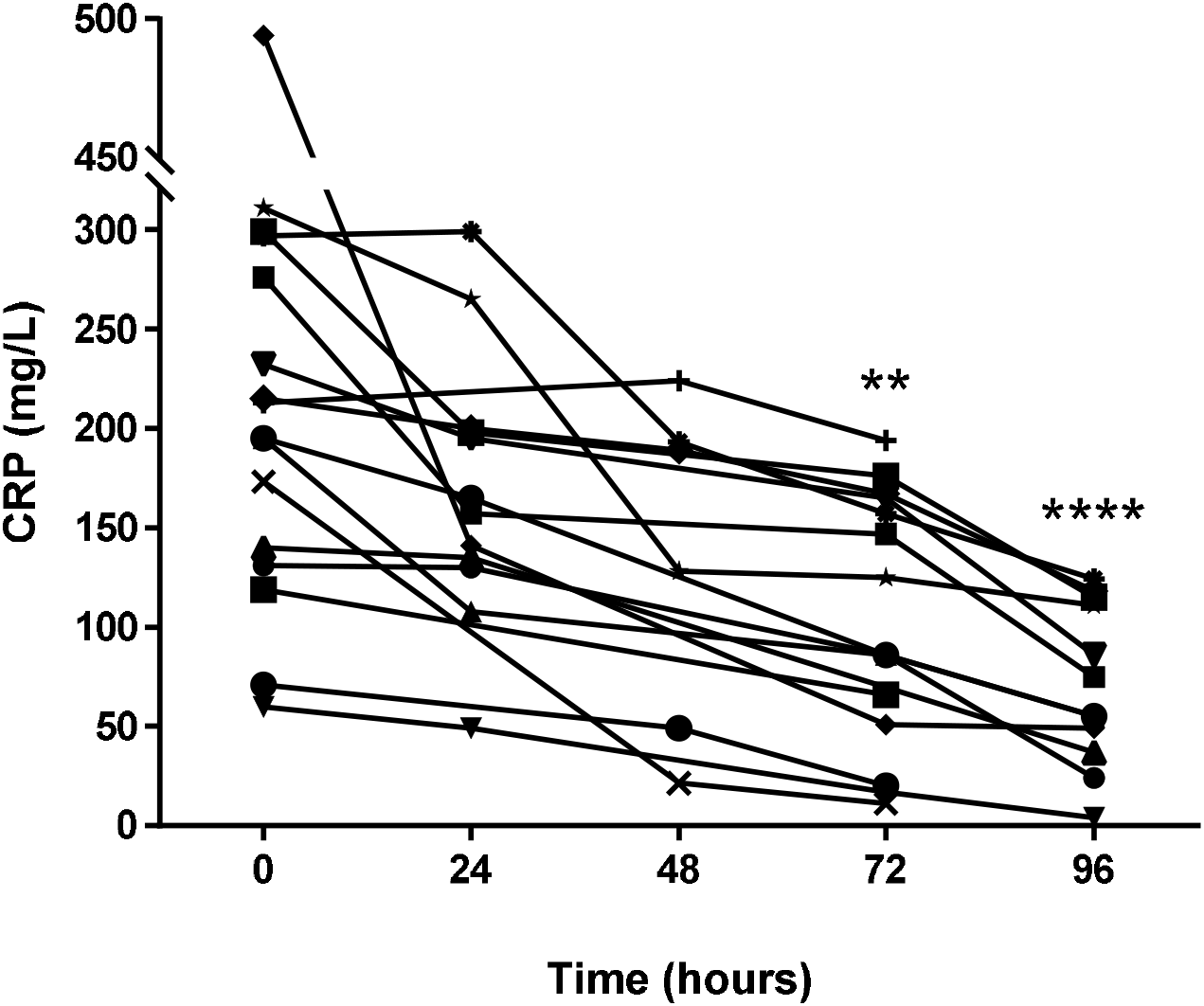
Effect of CIGB-258 on C-reactive protein (CRP) levels. Serum samples were obtained before treatment (0) and at 24, 48, 72 and 96 hours. Differences were analyzed using ANOVA and Tukey’s post-test (**P<0.001;**** P<0.0001)

IL-6 levels were investigated in the first seven patients included in this study. This cytokine was quantified in sera before treatment and at 24, 48, 72, and 96 hours after starting the therapy with CIGB-258. As shown in Figure 3, therapy with CIGB-258 led to a significant reduction of this cytokine.

**Figure 3.**
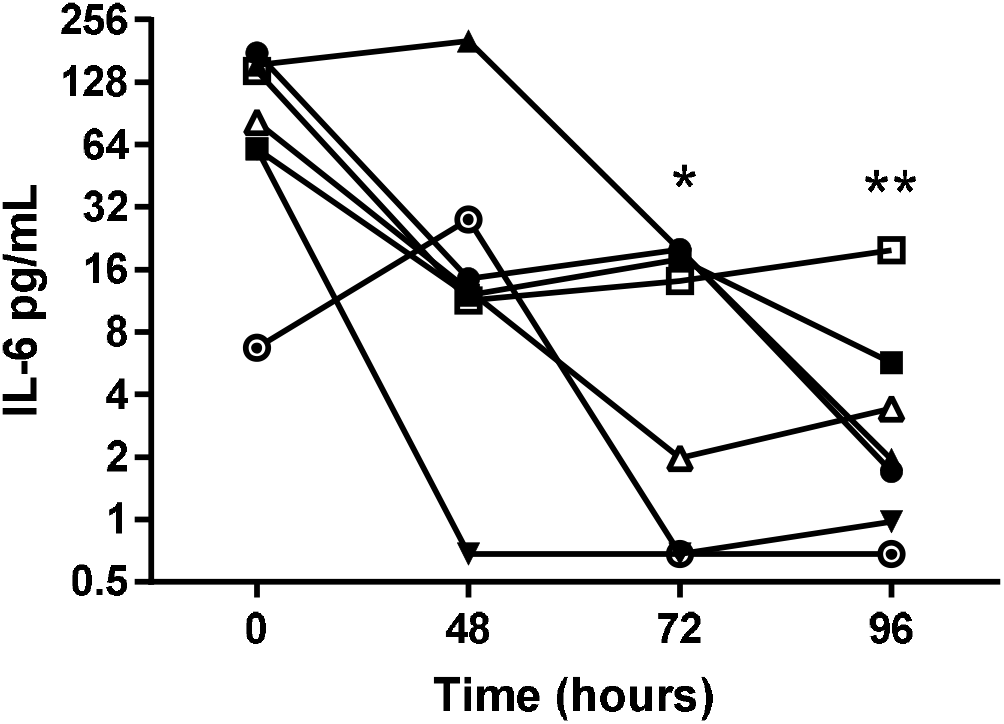
Effect of CIGB-258 on interleukin-6 (IL-6) levels. Serum samples were obtained before treatment (0) and at 24, 48, 72 and 96 hours. Differences were analyzed using Kruskal-Wallis and Dunn post-test (*P<0.05; **P<0,005)

### Safety

No adverse events associated with CIGB-258 were reported during therapy or the follow-up stage.

Additionally, before patients were discharged from the hospital they underwent a CT scan and no lesions associated with fibrotic events were found in their lungs.

## Discussion

Considerable evidence shows that a subgroup of patients with COVID-19 might have had a cytokine storm syndrome characterized by acute respiratory distress syndrome (ARDS) and secondary haemophagocytic lymphohistiocytosis (4).

Here, we report the effect of CIGB-258 therapy in seriously, or critically ill patients with COVID-19. These results supported the efficacy of CIGB-258 in the control of cytokine storms induced by SARSCoV-2.

CIGB-258 induced regulatory effects associated with the inhibition of inflammation in several experimental models and in RA patients (6,7,8,9).

A Phase I Clinical Trial with this peptide was carried out in 20 moderated active RA patients. Patients showed decreases in clinical activity of RA. CIGB-258 led to a significant reduction of interferon gamma (IFN-γ) and IL-17 (8).

In addition, this peptide induced a significant reduction of anti-CCP antibodies in patients that was related to clinical activity of RA (9).

A phase II clinical trial in RA patients is in progress, where 185 patients have been included. Therapy with this peptide in Phase I and II Clinical Trials has been safe for patients (8).

Recently, we studied the effect of CIGB-258 on PBMC isolated from RA patients by proteomic tools. This experiment showed that peptide administration induced a reduction of calprotectin levels (unpublished results). This protein is produced by monocytes and neutrophils. Calprotectin is an antimicrobial, calcium and zinc-binding heterocomplex protein contained in the cytosol fraction of innate immunity cells and released immediately after host-pathogen interaction. Thus, calprotectin has been proposed for the diagnosis of inflammatory conditions (10). Interestingly, the main biological pathway identified, that was associated with the molecular mechanism of CIGB-258, was NETosis (unpublished results).

All these findings indicate that CIGB-258 can consistently inhibit inflammation without causing immunosuppression. Under this rationale, we receive the approval of the Cuban Regulatory Authority (CECMED, http://www.cecmed.cu/) to start the use of CIGB-258 in the treatment of critically or seriously ill COVID-19 patients.

In this study, clinical and radiological improvement was observed in critically ill patients, after 48 hours of treatment with CIGB-258. Two patients even after recovering from respiratory distress and extubated had a fatal outcome due to nosocomial infections. These patients also received treatment with Itolizisumab, a CD-6 antigen specific monoclonal antibody (11).

In critically ill patients, gasometric parameters improved after the first administration of the peptide. These results are in correspondence with the pharmacokinetic and biodistribution of CIGB-258. Maximum concentration of CIGB-258 in the blood from rats was attained at 0.5 to 1 hour; and the t½ was calculated to be 6 hours, when the peptide was administered by the intravenous route.

Furthermore, the peptide had a wide biodistribution. CIGB-258 targets multiple organs including lungs (12). Monocytes, macrophages, and neutrophils are involved in the pathogenesis of respiratory distress. These cells secrete molecules that mediate inflammation (13).

Jingjiao et al in the first study to characterize PBMC proteome profile in mild and severe COVID-19 patients, identified nsp9 and nsp10 as potential virulence factors for SARS-CoV-2. These proteins interact with NKRF to facilitate IL-8/IL-6 induction, leading to uncontrolled infiltration and activation of neutrophils (14). According to our findings in proteomic studies CIGB-258 might, in fact, interfere with the activation of neutrophils and alveolar macrophage.

This hypothesis is in connection with the significant reduction of CRP and IL-6 levels induced by CIGB-258. IL-6 plays a significant role in inflammatory reaction and immune response. Several studies suggested that IL-6 is one of the most important cytokines involved in COVID-19-induced cytokine storms (4).

Unlike other treatments like Tocilizumab, Anankira and JAK kinases inhibitors, CIGB-258 does not produce immunosuppression.

Quantification of IL-6 in all patients, as well as other cytokines such as TNFa, IL-10 and calprotectin is ongoing.

Moreover, in preclinical models this peptide increased Treg. This effect may be important in the therapy of patients with cytokine storm, since Treg can contribute to the control of hyperinflammation. Furthermore, CIGB-258 therapy can be very effective in seriously ill patients.

Consequently, the five seriously ill cases were significantly improved. These patients did not require oxygen therapy after the first 24 hours of treatment with CIGB-258. Ferritin levels and D dimer gradually normalized after 48 hours of treatment. These preliminary results are encouraging and suggest that CIGB-258 has a broad spectrum of action on the inflammatory cascade including down-modulating active coagulation in COVID 19 patients.

Therapy with peptide was well tolerated in all patients. Patients discharged from the hospital are under strict pharmaco-surveillance.

CIGB-258 has proven to be an effective and safe treatment option for COVID-19 patients with cytokine storms. The early administration of CIGB-258 may improve seriously ill patients and avoid their progress to a critical condition.

CIGB-258 is currently being used in other Cuban hospitals for the treatment of seriously and critically ill COVID-19 patients. Studies to elucidate CIGB-258 mechanism of action are ongoing, but so far we can conclude that the application of CIGB-258 in COVID-19 patients effectively reduces the deterioration of their clinical condition offering an opportunity to avoid the fatal outcome of this infectious disease.

## Data Availability

The datasets used and/or analyzed during the current study available from the corresponding author on reasonable request

https://www.cigb.edu.cu/

## Acknowledgements

Study sponsorship was provided by CIGB and Ministry of Public Health of Cuba.

We thank the the nurses for their participation in the clinical work. We also thank Eduardo Pentón, Luis Javier Gonzalez, Marisol Cruz, Lourdes Hernandez, Hugo Nodarse, Claudia Urritia, Maria Vazquez, Amaury del Río, Yunaysy Jimenez and Ileana Delgado for their assistance and Hilda Garay, Ever Perez and Matilde Lopez for their contribution in the drug production process and formulation.

## Authors’ contributions

VRR was the principal investigator, participated in trial design and execution, performed data review and reviewed the manuscript. SSR and PRR participated in trial execution. BRM and HCMperformed IL-6 data analysis, drafted figures and tables and reviewed the manuscript. SLB;GA;APR;OPD and OGLM participated in trial execution. Baldomero JE collects and reviews patient’s data. CLR adviser trial execution,collects and reviews patient’s data. GG participated in trial design, drafted the manuscript. MDG is manager project anddrafted the manuscript. DHMC generated the molecule under study, projectleader, served as scientific advisor and drafted the manuscript. All authors revised and approved the final version of this manuscript.

